# Projection of COVID-19 Cases and Deaths in the US as Individual States Re-open May 4, 2020

**DOI:** 10.1101/2020.05.04.20090670

**Authors:** Teresa Yamana, Sen Pei, Sasikiran Kandula, Jeffrey Shaman

**Affiliations:** Department of Environmental Health Sciences, Mailman School of Public Health, Columbia University

## Abstract

In March and April 2020, control measures enforcing social distancing and restricting individual movement and contact were adopted across the United States in an effort to slow the spread and growth of COVID-19. However, a number of states have now begun to ease these restrictions. Here, we evaluate the effects of loosening stay-at-home orders on COVID-19 incidence and related outcomes. We use a metapopulation model applied at county resolution to simulate the spread and growth of COVID-19 incidence in the United States. We calibrate the model against county-level daily case and death data collected from February 21, 2020 to May 2, 2020, and project the outbreak in 3,142 US counties for 6 weeks into the future. Projections for daily reported cases, daily new infections (both reported and unreported), new and cumulative hospital bed demand, ICU bed and ventilator demand, as well as daily mortality, are generated. We observe a rebound in COVID-19 incidence and deaths beginning in late May, approximately 2 to 4 weeks after states begin to reopen. Importantly, the lag between infection acquisition and case confirmation, coupled with insufficient broader testing and contact tracing, will mask any rebound and exponential growth of the COVID-19 until it is well underway.

## Introduction

Twenty-five states have or plan to partially re-open their economies in the coming week. The impact of this increased activity on contact patterns, the transmission of SARS-CoV-2 and COVID-19 incidence remains highly uncertain, as levels of compliance with social distancing, return to work, and consumer willingness to frequent businesses are unknown. While it is certainly possible that strong social distancing practices enforced in stores, restaurants and theaters, as well as increased use of face masks, could offset any increases in person-to-person contact, congregation in common spaces and associated increased opportunities for virus transmission, it is likely that a return to greater activity will reverse some of the gains, i.e. reductions of virus transmissibility, accrued over the last 6 weeks, particularly if initial re-openings do not produce an immediate growth of cases and consumer confidence grows.

Here we project the effects of weak increases of transmissibility, relative to current estimates of effective reproduction number, *R_eff_(t)*, on COVID-19 outcomes over the next 6 weeks. Increased virus transmissibility is only applied to states in which a re-opening of the economy has or is slated to occur.

## Projection scenarios

Projections are generated using a county-scale metapopulation model^1^ optimized to daily confirmed COVID-19 cases and deaths from February 21 – May 2, 2020. The model optimization process, described below, results in county specific parameter estimates. We then use these parameter values as starting points for projecting future disease transmission. In Pei and Shaman^1^ we implemented 3 hypothetical control scenarios to reflect reduced contact between individuals due to stay-at-home orders, as well as reactive personal decisions to limit social contacts: starting from 14 days prior to the projection day, we applied 20%, 30% and 40% weekly reductions of the contact rate *(β)* in counties with ≥10 reported cases in a week, until the weekly numbers of new confirmed case in those counties decreased (i.e., the curve flattened).

Here, we modify our control scenarios to account for increases in contact rates due to loosening restrictions in states that have begun to reopen economically. We project three scenarios. In the first two scenarios, for states maintaining or increasing current social distancing restrictions, we continue to apply the 20% weekly reductions of contact rates, as described above. However, in states that reopen, we apply an increase to the contact rate of counties in those states. In the first scenario, we apply a one-time 10% increase to the contact rate during the week that the state is scheduled to reopen and maintain this new increased rate for the remainder of the projection. In the second scenario, the contact rate is increased by an additional 10% each week to represent progressive loosening of restrictions and increased public confidence and frequenting of businesses. A third comparison scenario assumes no effect due to reopening, and instead applies 20% weekly reductions in contact rates for all counties meeting the criteria described above, and as in Pei and Shaman^1^. These 3 scenarios are summarized as follows:

1. Weekly 20% decrease in places with growing weekly cases and a one-time 10% increase in places with return to work (latter supersedes the former)
2. Weekly 20% decrease in places with growing weekly cases and a weekly 10% increase in places with return to work (latter supersedes the former)
3. Weekly 20% decrease in places with growing weekly cases

The state-by-state reopening schedules used in our projections are shown in Table 1. We do not differentiate between the types of business and activities allowed to reopen in each state. We also do not differentiate within a state; increased contact rates are applied to all counties in a state with loosening restrictions.

**Table 1.** Reopening schedule as of 5/02/2020

| **state** | **date** |
| --- | --- |
| South Carolina | 4/20/20 |
| Alaska | 4/24/20 |
| Oklahoma | 4/24/20 |
| Montana | 4/26/20 |
| Colorado | 4/27/20 |
| Georgia | 4/27/20 |
| Tennessee | 4/27/20 |
| Mississippi | 4/28/20 |
| South Dakota | 4/28/20 |
| Alabama | 5/1/20 |
| Idaho | 5/1/20 |
| Iowa | 5/1/20 |
| Maine | 5/1/20 |
| North Dakota | 5/1/20 |
| Texas | 5/1/20 |
| Utah | 5/1/20 |
| Wyoming | 5/1/20 |
| Indiana | 5/2/20 |
| Florida | 5/4/20 |
| Kansas | 5/4/20 |
| Minnesota | 5/4/20 |
| Missouri | 5/4/20 |
| Nebraska | 5/4/20 |
| Ohio | 5/4/20 |
| West Virginia | 5/4/20 |

## Model

We use a metapopulation SEIR model^1^ to simulate the transmission of COVID-19 among 3,142 US counties. This model has been described previously in Pei and Shaman^1^ and has been used since March 13, 2020 to make biweekly projections of COVID-19 incidence, hospitalizations and deaths. The model represents two types of movement: daily work commuting and random movement. Information on county-to-county work commuting is publicly available from the US Census Bureau^2^, which is used to determine rates of intercounty movement prior to March 15, 2020. We assume the number of random visitors between two counties is proportional to the average number of commuters between them. After March 15, 2020, we use SafeGraph estimates of the reduction of inter-county visitor numbers in points of interest (POI) (e.g., restaurants, stores, etc.) to inform the decline of inter-county movement on a county-by-county basis. For instance, if the number of out-of-county visitors was reduced by 30% in a county on a given day comparing with the baseline on March 15, 2020, the size of subpopulations traveling to this county would be reduced by 30% accordingly. During projection into the future, we maintained the most recent level of inter-county movement.

As population present in each county is different during daytime and nighttime, we model the transmission dynamics of COVID-19 separately for these two time periods. We formulate the transmission as a discrete Markov process during both day and night times. The daytime transmission lasts for *dt*_1_ day and the nighttime transmission *dt*_2_ day (*dt*_1_ + *dt*_2_ = 1). Here, we assume daytime transmission lasts for 8 hours and nighttime transmission lasts for 16 hours, i.e., *dt*_1_ = 1/3 and *dt*_2_ = 2/3. The transmission dynamics are depicted by the following equations.

Daytime transmission:

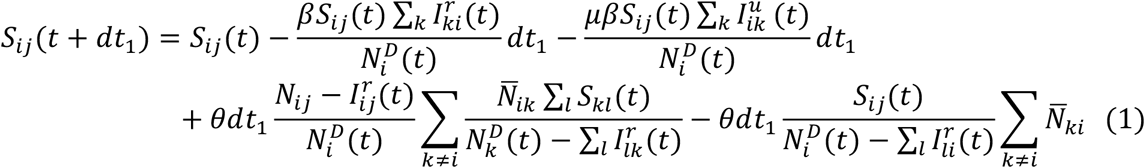

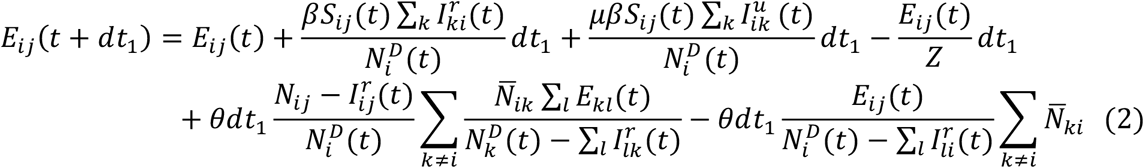

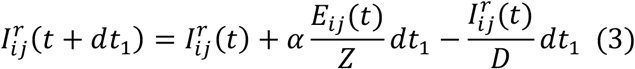

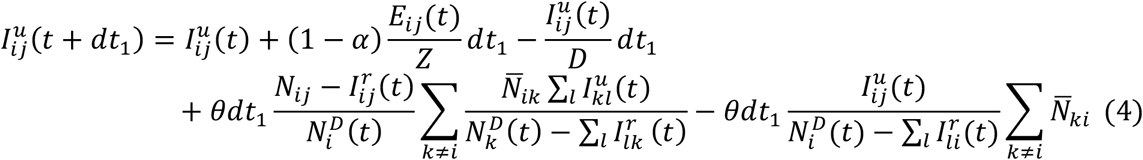

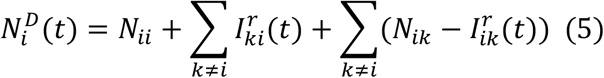

Nighttime transmission:

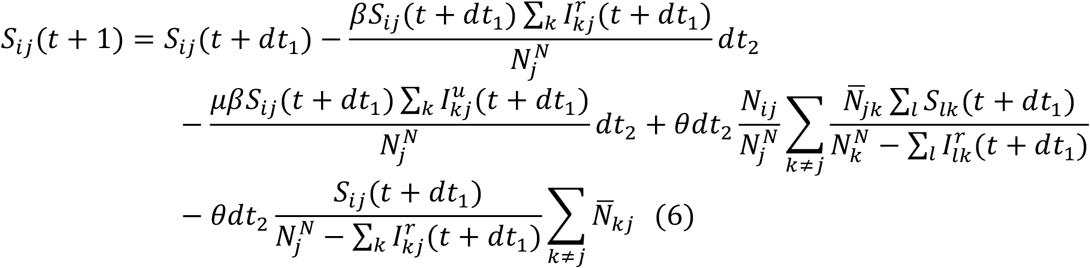

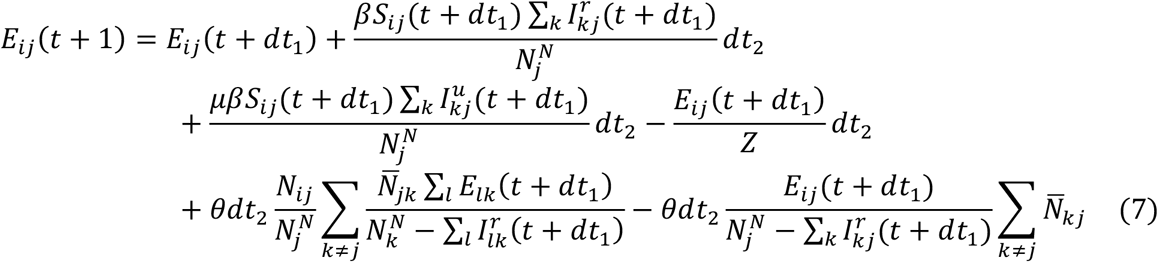

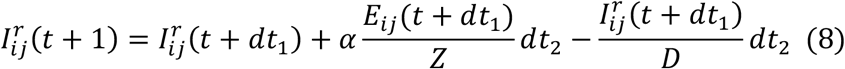

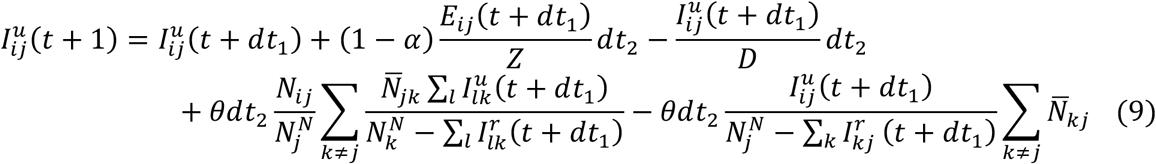

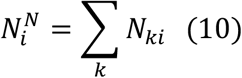

Here, *S_ij_, E_ij_*, 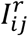, 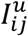 and *N_ij_* are the susceptible, exposed, reported infected, unreported infected and total populations in the subpopulation commuting from county *j* to county *i (i ← j*); *β* is the transmission rate of reported infections; *μ* is the relative transmissibility of unreported infections; *Z* is the average latency period (from infection to contagiousness); *D* is the average duration of contagiousness; *α* is the fraction of documented infections; *θ* is a multiplicative factor adjusting random movement; 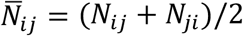 is the average number of commuters between counties *i* and *j;* and 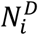 and 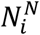 are the daytime and nighttime populations of county *i*. We integrate Eqs. 1-10 using a Poisson process to represent the stochasticity of the transmission process. A similar model has been used to generate forecasts for influenza in the United States^3^.

To account for reporting delay, we mapped simulated documented infections to confirmed cases using a separate observational delay model. In this delay model, we account for the time interval between a person transitioning from latent to contagious (i.e. 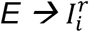) and observational confirmation of that individual infection. To estimate this delay period, *T_d_*, we examined line-list data from early-confirmed cases in China^4^. Prior to January 23, 2020, the time-to-event distribution of the interval (in days) from symptom onset to confirmation is well fit by a Gamma distribution^5^ *(a =* 1.85, *b =* 3.57, *LL =* -252.24). Consequently, we adopted a Gamma distribution to model *T_d_*, but tested longer mean periods (*ab*) as symptom onset often lags the onset of contagiousness. Our analysis indicates that an average delay of 9 days supports better fitting to the daily incidence data. As a result, we adopted *T_d_* = 9 days in the projection.

To map the simulated infections (both documented and undocumented) to death, we used the age-stratified infection fatality rate (IFR) reported in Verity et al^6^. The IFR in each county was computed as a weighted average using demographic information on age structure. Based on observations for the US, the national death curve has a 7-day lag with respect to the incidence curve. As a result, we used a gamma distribution with a mean of 16 days *(a =* 1.85) to represent the delay between a person transitioning from latent to contagious and death.

We used the hospitalization data in COVID-NET^7^ to estimate rates of hospitalization for confirmed cases. State-level COVID-19 hospitalization rates are available for 14 states. Applying these rates to case data, we computed the percentage of COVID-19 confirmed patients that are hospitalized in these states. For states without hospitalization data, the national average value was used. The county-level hospitalization rate for confirmed cases is defined using state-level estimate. We modeled the time from contagiousness to hospitalization using a gamma distribution (*T_d_*=6 days, a=4, b=1.5), based on information from New York City.

### Estimate parameters on March 13 2020

To derive an estimate of prior parameters on March 13 2020, we calibrated the transmission model against county-level incidence data reported from February 21, 2020 through March 13, 2020^8^. Specifically, we estimated model parameters using an iterated filtering (IF) framework^9,10^. The metapopulation model is high dimensional with 60,232 subpopulations. We therefore applied an efficient data assimilation algorithm - the Ensemble Adjustment Kalman Filter (EAKF)^11^, which is applicable to high dimensional model structures, in multiple iterations to infer parameters *β, μ*, *Z, D, α* and *θ*. This iterated filtering (IF)-EAKF framework has previously been used to infer parameters in a large-scale agent-based model for antimicrobial-resistant pathogens^12^, as well as a metapopulation model depicting the spread of SARS-CoV-2 in China^5^. Details of its implementation can be found in Ref. 5.

The prior ranges of model parameters were set as: *β ∈* [0.3,1.5], *μ ∈* [0.2,1.0], *Z ∈* [2,6], *D* ∈ [2,6], *α* ∈ [0.02,1.0], and *θ ∈* [0.01,0.3]. In the inference, we fixed the shape parameter of the Gamma distribution for *T_d_* as *a =* 1.85, and vary the mean value of the distribution. We tested a range of mean *T_d_* values from 6 days to 10 days, and found that *T_d_* = 9 days supports the best fitting.

To initialize the model, we seeded exposed individuals *(E)* and unreported infections (*I^u^*) in counties with at least one confirmed case. Unlike the situation in China, where the outbreak originated from a single city, there was importation to multiple locations in the US that could have initiated community transmission. To reflect this potential ongoing community transmission before the reporting of the first local infection, for each county with confirmed cases, we randomly drew *E* and *I^u^* from uniform distributions [0, 12*C*] and [0, 10*C*] 8 days prior to the reporting date *(T*_0_) of the first case. Here *C* is the total number of reported cases between day *T*_0_ and *T*_0_ *+* 4.

The rationale for this seeding strategy is as follows. If an average reporting delay of 8 days is assumed, we can estimate *I^r^* on day *T*_0_ − 8 by 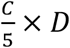, where 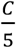 is the average number of daily cases during the first five days with reporting *(T*_0_ to *T*_0_ *+* 4). If we use the upper bound of the prior for D (i.e., 5 days), *I^r^* is estimated as *C*, which is also an upper bound. Using parameters obtained from China^1^, we assume the mean *I^u^* on day *T*_0_ *−* 8 is 5*C*, implying a reporting rate of 1/6=16.7%. Drawing *I^u^* from [0,10*C*] leads to a broader prior range of the reporting rate. As both *I^r^* and *I^u^* were evolved from the exposed population *E*, we draw *E* from the range [0,12*C*]. This crude calculation gives an estimate of seeding in US counties. During inference, this seeding can be adjusted up or down by the filter, and best-fitting models produce simulations that capture observed outcomes.

### Model calibration after March 13 2020

In order to represent variability in contact rates and reporting rates through space and time, starting from March 13, we introduced separate estimates for *β* and *α* in each county. The prior ranges for the other model parameters were taken from the posterior estimate for March 13, 2020 as described above.

We defined a separate set of parameters *α_i_* and *β_i_* for each county, which were estimated by the EAKF for each county after March 13, 2020. The prior contact rates in all counties were scaled based on their population density using the following relation:

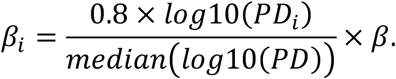

Here *PD_i_* is the population density in county *i, median(log*10*(PD))* is the median value of log-transformed population density among all counties, and *β* is the contact rate estimated before March 13 2020.

From March 13 2020 through May 2, 2020, we performed EAKF inference each day using both incidence and death data. To account for reporting delays of confirmed cases and deaths, we used incidence and death numbers 6 days ahead to constrain model states variables and parameters. The latest estimates of parameters (i.e. for May 2, 2020) were then used to generate projections for 6 weeks onwards.

## Results

The estimated values of the effective reproductive number (R_eff_) as of May 2, 2020 are shown in Figure 1 for counties in states with scheduled re-openings. Many counties are near or above 1, which is the value above which we expect to observe increasing infections. As such, even a small increase in the contact rate can drive *R*_eff_ greater than 1, which would then lead to a rebound in transmission and a lagged increase of confirmed cases, hospitalizations and deaths. These findings indicate that most states are not well-positioned to re-open their economies and simultaneously control the spread of COVID-19 infections.

**Figure 1:**
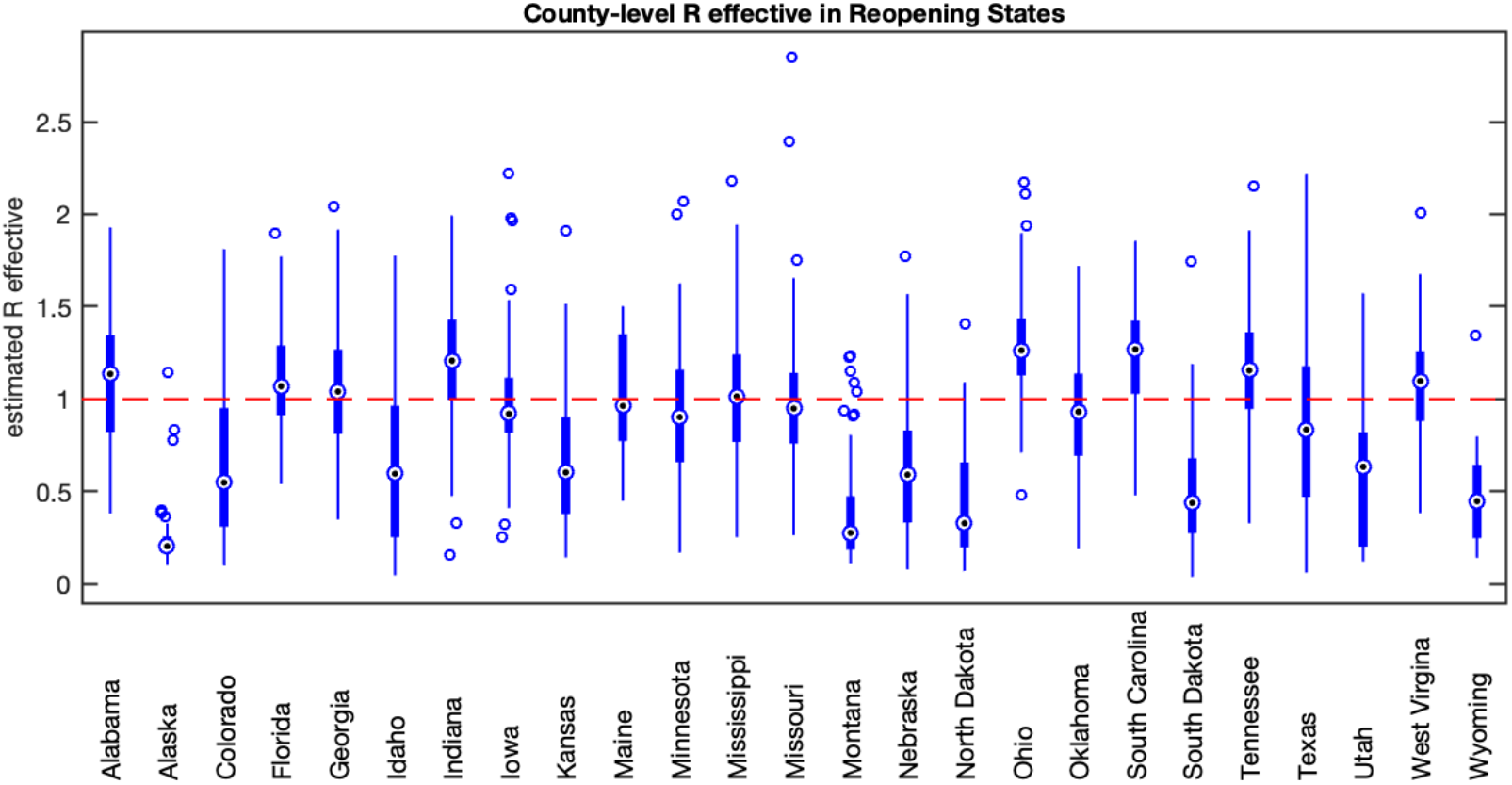
Effective reproduction number for counties in reopening states as of May 2, 2020. Each data point is the mean estimated *R*_eff_ for each county within the state. Boxes show *R*_eff_ for the median, 25^th^ and 75^th^ percentile counties. The red line indicates an effective reproduction number of 1.

Both scenarios with increasing contact rates in reopening states resulted in a rebound in COVID-19 incidence, hospitalizations, and deaths at the national scale (Figure 2). The rebound was faster and stronger for the weekly-increase scenario. Notably, the increase in cases and deaths is not apparent at the national scale until two to four weeks after the first states begin to reopen.

**Figure 2:**
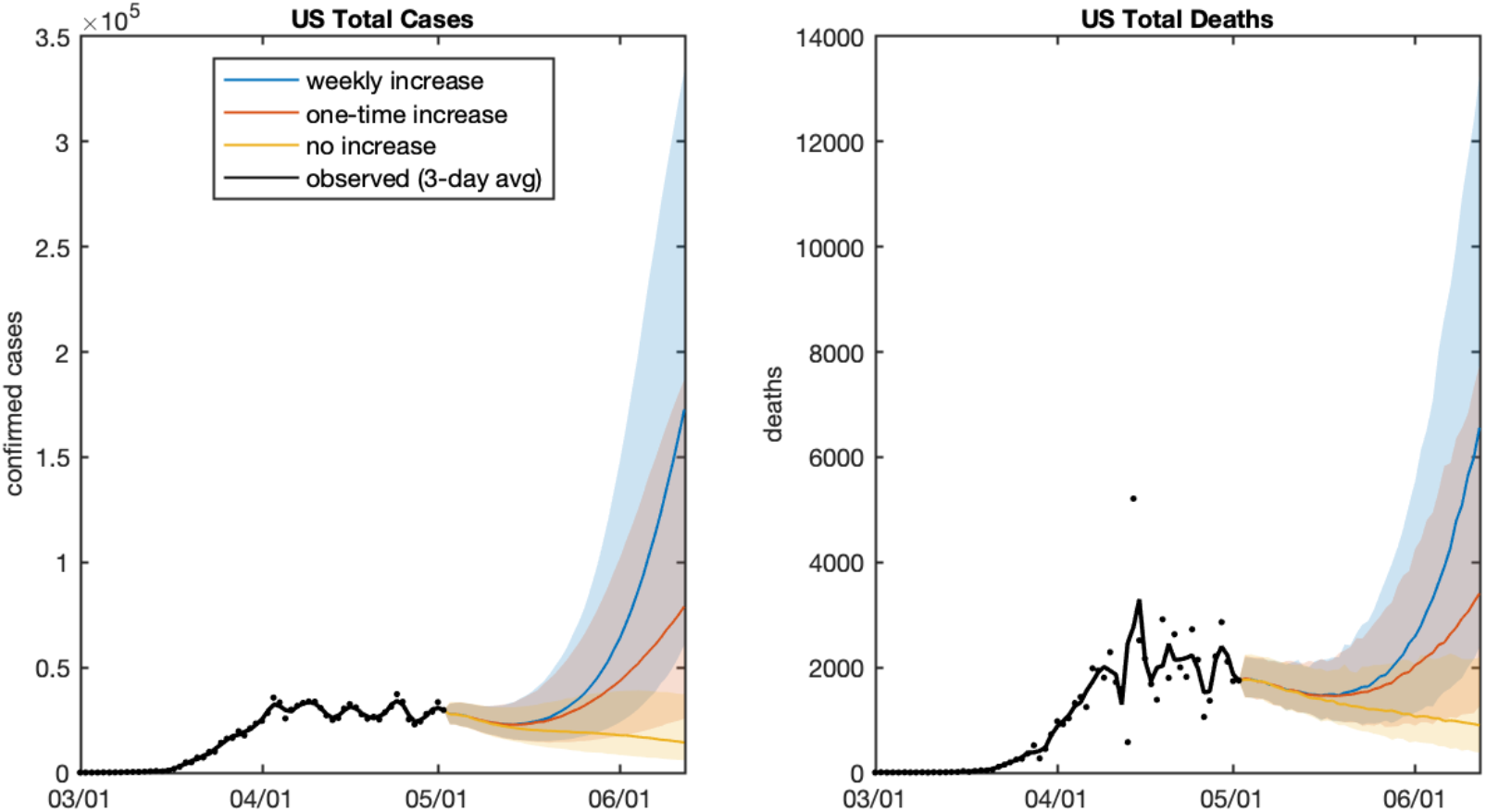
Effect of reopening states in 20% reduction scenario. Daily projected and observed new cases, hospitalizations and deaths. The shaded regions show the 95% credible intervals.

Projections of COVID-19 incidence are shown in Supplemental Figures 1 and 2 for reopening states, and Supplemental Figures 3 and 4 for states that remain closed. With few exceptions, reopening states are projected to experience exponential growth of both cases and deaths; the exceptions correspond with the states with the lowest values of *R*_eff_ (Figure 1). States with restrictions remaining in place are projected to have decreasing or stable numbers of cases and deaths under all three scenarios.

The projection results are available at GitHub: https://github.com/shaman-lab/COVID-19Projection.

### Interpretation Considerations

Several qualifications with respect to these projections must be noted and considered during interpretation. Firstly, the model is optimized using observations through May 2, 2020; however, those observations, i.e. confirmed cases and deaths by county, represent infections that were acquired by individuals 1-3 weeks earlier. Because of this long delay, the effects of changes in social distancing and contact patterns over the last 3 weeks on virus transmission have yet to be fully observed.

Secondly, the landscape to which this model has been optimized is highly variable in space and time, due to differences in contact behavior, population density, control measures and testing practices. These differences in space and time make the fitting of any model of this scale challenging.

Finally, we expect that the response to COVID-19 transmission will be adaptive at both the government and the individual level. We expect that if states begin to experience a rise in infections and deaths as shown in these projections (following a 2-4 week delay), further restrictions on contacts will be put into place to counter such a trajectory.

## Data Availability

All input data are publicly available; the model output is available at https://github.com/shaman-lab/COVID-19Projection

---

**Supplemental Figure 1.**
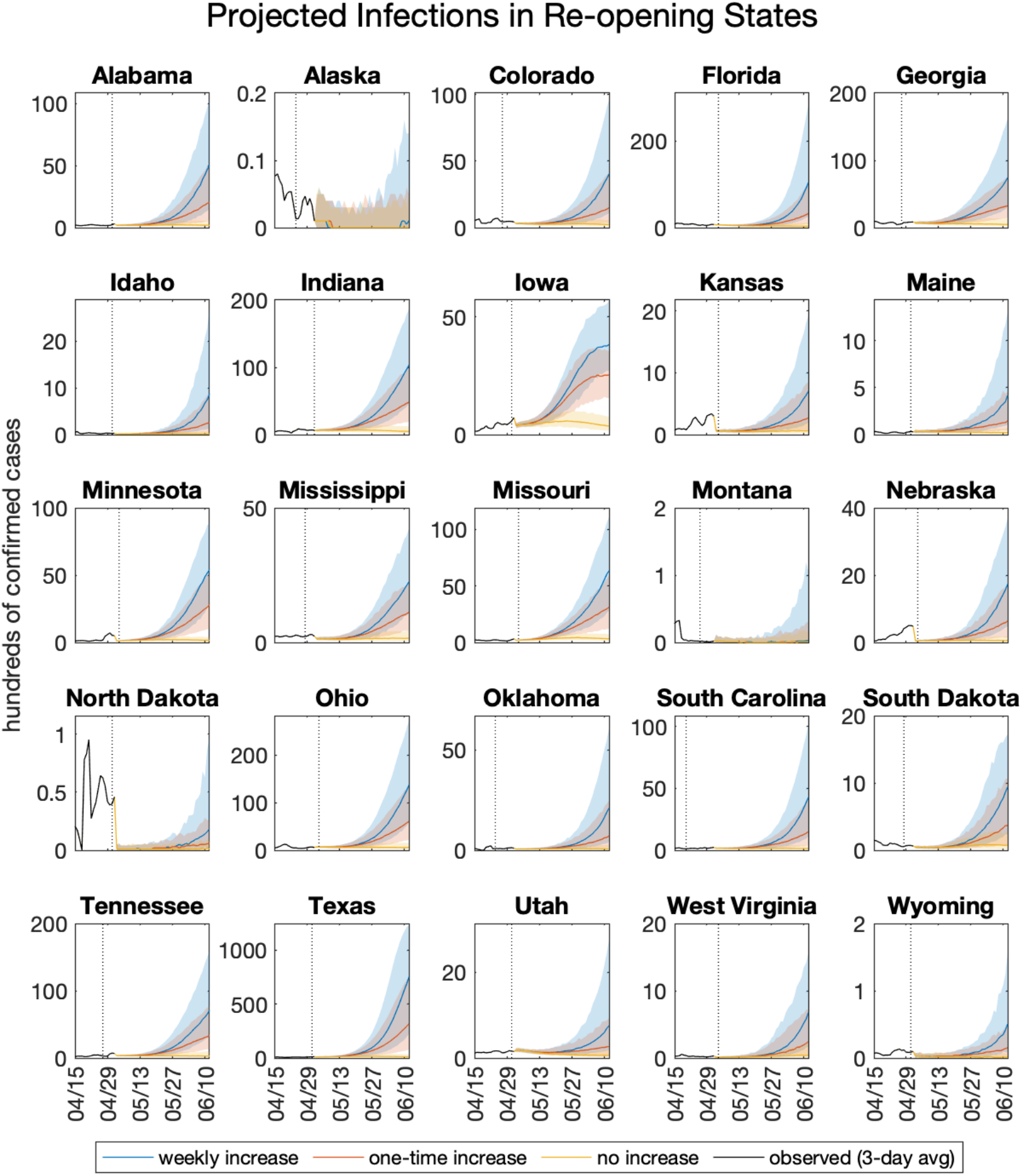
Projected infections in states with restrictions remaining in place. The dashed vertical line indicates the date of reopening. Shaded regions indicate 95% credible intervals.

**Supplemental Figure 2.**
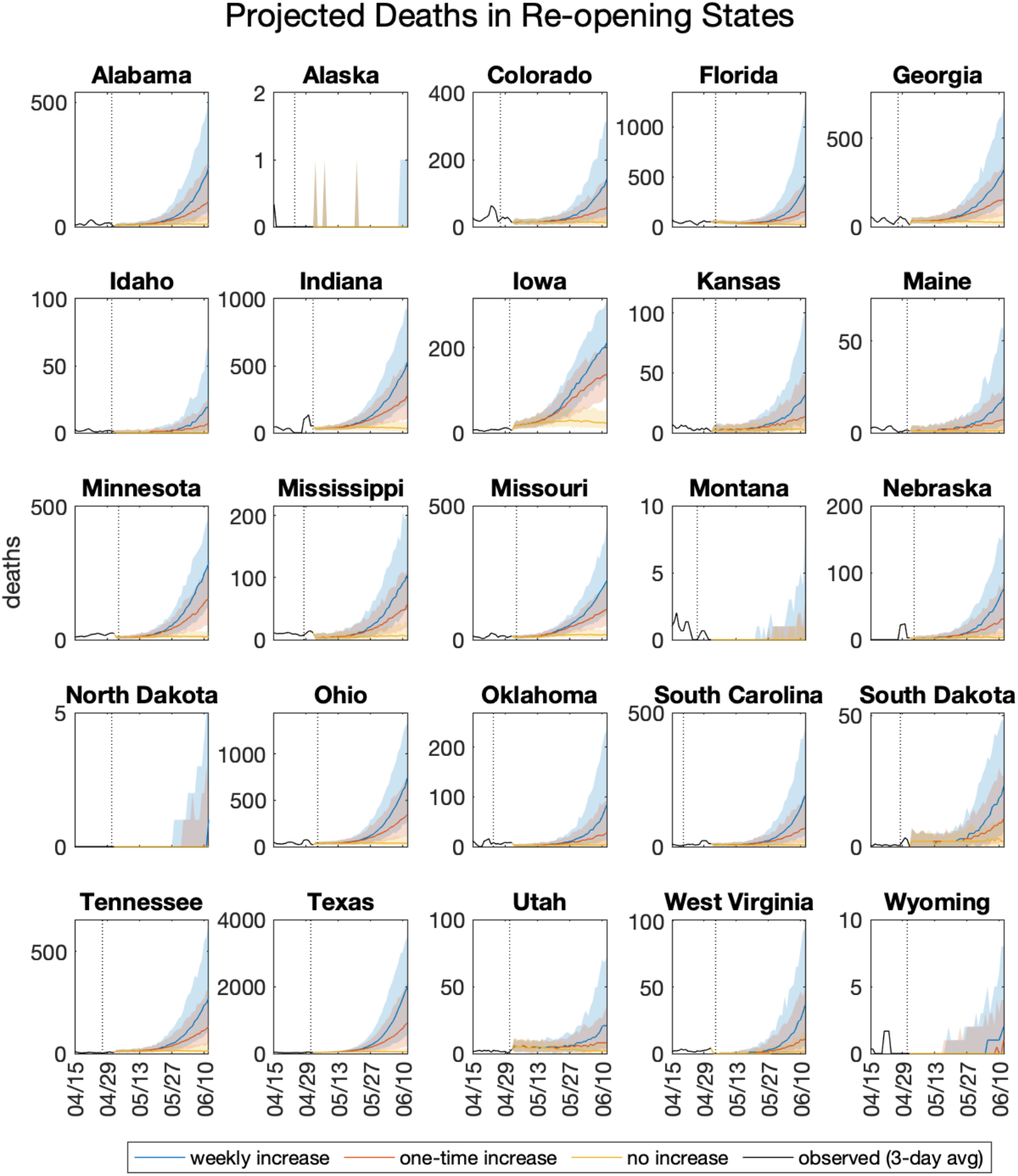
Projected deaths in states with restrictions remaining in place. The dashed vertical line indicates the date of reopening. Shaded regions indicated 95% credible intervals.

**Supplemental Figure 3.**
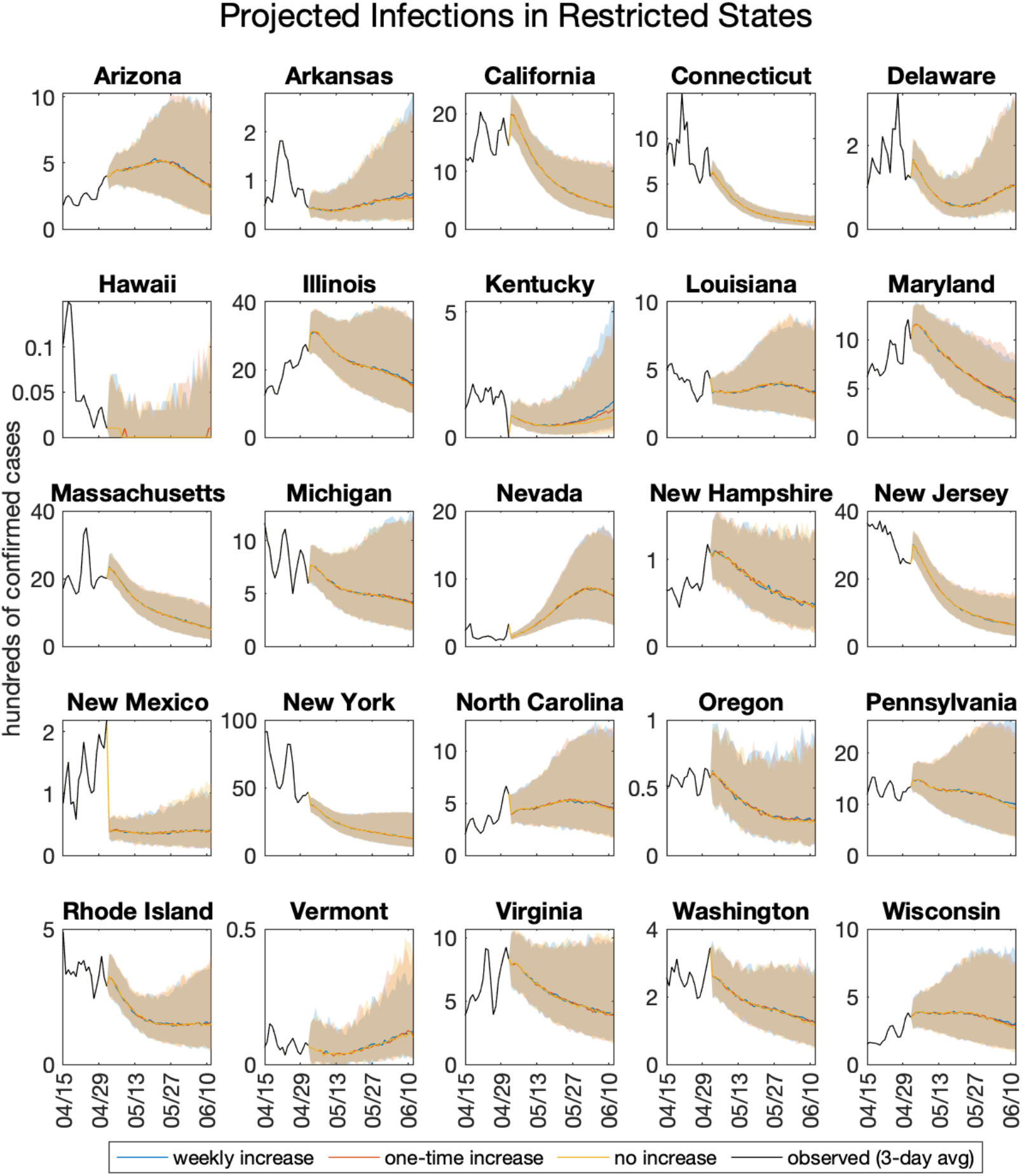
Projected infections in states with restrictions remaining in place. Shaded regions indicate 95% credible intervals. The three scenarios result in overlapping projections, as no increase in contact rate is applied to these states.

**Supplemental Figure 4.**
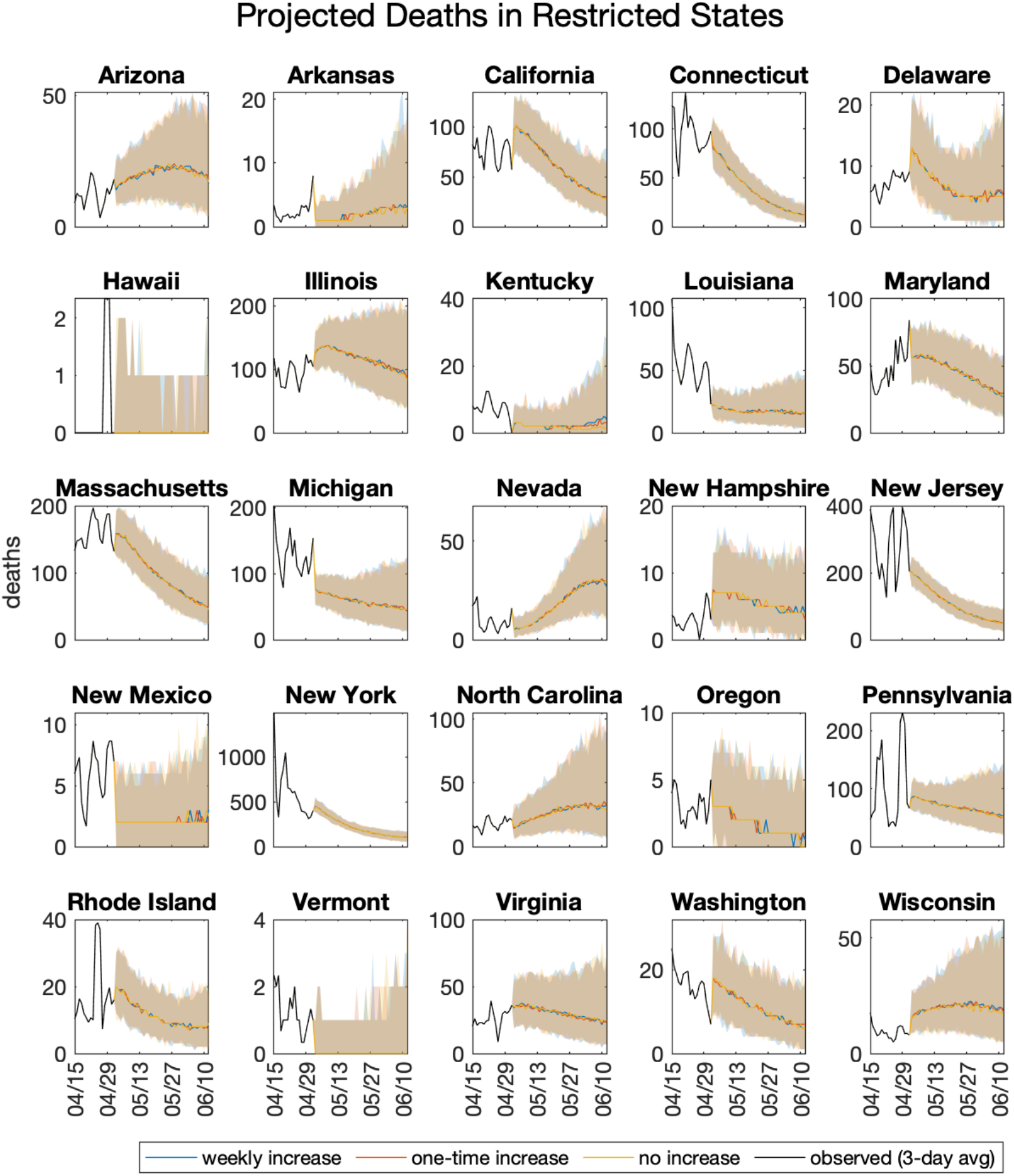
Projected deaths in states with restrictions remaining in place. Shaded regions indicate 95% credible intervals. The three scenarios result in overlapping projections, as no increase in contact rate is applied to these states.

